# The Inherited Retinal Disease Pathway in the United Kingdom: a Patient Perspective and the Potential of AI

**DOI:** 10.1101/2025.01.14.25320497

**Authors:** Wendy Wong, Dayyanah Sumodhee, Tiyi Morris, Bhavna Tailor, Catherine Hollyhead, William A Woof, Stephen Archer, Carl Veal, Loy Lobo, Saoud Al-Khuzaei, Malena Daich Varela, Thales Antonio Cabral De Guimaraes, Manuel Gomes, Mital Shah, Susan M Downes, Savita Madhusudhan, Omar A Mahroo, Andrew R Webster, Michel Michaelides, Nikolas Pontikos

## Abstract

**Background:** Inherited Retinal Diseases (IRDs) are the leading cause of blindness in young people in the UK. Despite significant improvements in genomics medicine, diagnosis of these conditions remains challenging, with many patients enduring lengthy diagnostic odysseys and even after genetic testing around 40% of them do not receive a definite genetic diagnosis. This survey aims to explore the experience of individuals affected with IRDs, their relatives, friends and caregivers, and the potential acceptability of an AI technology, such as Eye2Gene.

**Methods:** This cross-sectional survey was distributed electronically using the Qualtrics-encrypted platform between April to August 2024. The mixed-methods survey included Likert-scale and open-ended queries. Analysis was performed using descriptive statistics and content methods.

**Results:** The survey was answered by 247 respondents of which 79.8% were patients and the remainder were relatives, friends and caregivers. There was substantial variability in patient diagnostic journeys in terms of waiting times to see a specialist (IQR 1 to 4 years), commute required (IQR 10 to 74 miles) and number of visits to reach a diagnosis (IQR 2 to 4). A substantial proportion of patients had a change in diagnosis had a change in diagnosis (35.8%). The majority of respondents were overwhelmingly in favour of the integration of AI into the IRD pathway to accelerate genetic diagnosis care (>90%).

**Conclusion:** This survey identifies several key gaps and disparities in the IRD pathway which can be addressed in part by the integration of AI for more equitable care. Survey also revealed a favourable attitude towards incorporating AI into the diagnostic testing of IRDs.

**Synopsis:** A survey by 247 people directly or indirectly affected by inherited retinal diseases in the UK reports substantial gaps and disparities in the patient diagnostic pathway which could in part be addressed by Artificial Intelligence.

## Introduction

Inherited retinal diseases (IRDs) are a significant cause of irreversible vision loss, affecting up to one in 1380 individuals worldwide, and represent a leading cause of blindness in the working-age population and children [1–4]. The term IRD encompasses a large group of genetically predetermined retinal conditions that have been linked to pathogenic variants in over 300 different gene variants [5,6]. The International Rare Diseases Research Consortium (IRDiRC) has set a commendable target of reducing the diagnostic delay for known medical disorders to under one year, but the average time to diagnosis for an IRD patient in Europe or America is presently 6.4 ± 9.1 years [7,8]. Reaching a definitive diagnosis for patients with IRDs within the significant genetic heterogeneity, the pronounced phenotypic variability, and the presence of modifier loci as well as limited available clinical expertise for IRDs [9].

Although next-generation sequencing technology offers a powerful strategy to shorten the diagnostic process for patients with IRD, a genetic diagnosis is still not obtained in 26–48% of individuals [10]. The bioinformatics pipeline has been implicated as one of the major sources of variability in variant detection [11]. Another challenge lies in interpreting novel or rare genetic variants, which can yield an inconclusive result as a variant of uncertain significance. Some of this ambiguity can be resolved where the patient’s phenotype is highly specific for the disease and can be used to support the pathogenicity of the detected variants [12] The importance of evaluating the phenotype in variant interpretation is also evident from the guidelines jointly published by the American College of Medical Genetics and Genomics and the Association for Molecular Pathology, stating that ‘clinical laboratories are encouraged to form collaborations with clinicians to provide clinical information to better understand how genotype influences clinical phenotype and resolve differences in variant interpretation [12].

Multi-modal imaging techniques are an important tool in obtaining phenotypic information and valuable diagnostic markers when diagnosing IRD[13]. The significant processing power and speed of currently available artificial intelligence (AI) based techniques can link large volumes of multimodal imaging data acquired from patients with IRDs enabling identification of distinct patterns of neuroretinal degeneration and genotype-phenotype correlations [14,15]. These developments in AI technology can potentially aid in accelerating the diagnostic process while also providing more equitable access to diagnostic expertise for patients with IRDs. Moreover, AI analysis can potentially help in interpreting the genetic results and could aid in obtaining a molecularly confirmed diagnosis in the currently ‘‘unsolved cases’’. This is of significant importance as obtaining a genetic diagnosis helps provide information on the inheritance pattern, supports family planning decisions, enables clinicians to provide prognostic information, facilitates enrolment to disease registries, and is required for recruitment to clinical trials [16].

## Methods

### Survey design

The survey was designed and iteratively refined through a three-stage process [17]. The first stage involved an expert working group arising from the collaboration between a bioinformatician specialised in artificial intelligence (N.P), a health psychologist (D.S) and ophthalmologists specialising in IRDs (A.R.W, M.M and O.A.M). The second stage included patient advisory groups (B.T, C.H, S.A, C.V and L.L) to obtain feedback on the clarity of the questions, ensure the survey adequately captured the patient perspective and that all questions were framed with appropriate sensitivity. The final stage was a pilot to identify any areas of improvement and to obtain an estimate of the time required to complete the survey.

The survey was designed on Qualtrics (Provo, UT), which dynamically presents the next question based on the submitted response. This adaptive approach was chosen to reduce the burden on respondents by only showing relevant questions and to increase the likelihood of the survey participants completing the survey in its entirety. The survey included a mix of closed- and open-ended questions. A Likert scale was used for opinion-related questions. There was a total of 8 main questions, each with multiple subparts (supplementary information).

This research was approved by the IRB and the UK Health Research Authority Research Ethics Committee (REC) reference (22/WA/0049) “Eye2Gene: accelerating the diagnosis of inherited retinal diseases” Integrated Research Application System (IRAS) (project ID: 242050). All research adhered to the tenets of the Declaration of Helsinki.

Consent was obtained at the start of the survey. Respondents that did not select the option for consent to participate in the survey were excluded.

### Survey distribution

The survey was distributed electronically using an online secure platform (Qualtrics), through the email distribution lists of registered charities in the United Kingdom which provide support to patients with IRDs: Retina UK, Moorfields Eye Charity and Stargardt Connected, to include individuals affected by IRDs, their relatives, friends and caregivers.

### Data analysis

Data were downloaded from Qualtrics exported into a Microsoft Excel file and examined for duplicate responses by checking if the internet protocol (IP) address indicated multiple submissions from the same survey respondent. After excluding duplicate responses, quantitative responses were analysed using pivot tables in Excel.

## Results

Between 22^nd^ April 2024 and 12^th^ Aug 2024, 247 fully completed responses were received. Five responses were excluded from the analysis: 4 were duplicates and 1 respondent did not provide consent to participate.

### Demographic Characteristics of the Survey Respondents

The majority of respondents (79.8%) were patients; the remaining were relatives, caregivers or close friends (**Table 1**). The median (interquartile range) age of the participants was 53 (36-64) years and 57.4% were female. The most common conditions were Stargardt disease (44.2%) and retinitis pigmentosa (RP) (43.0%).

**Table 1.**
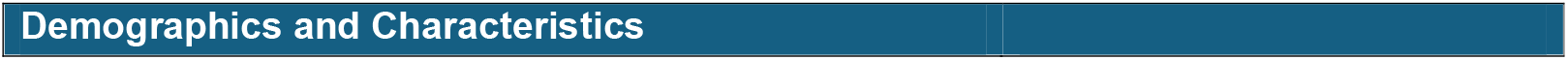
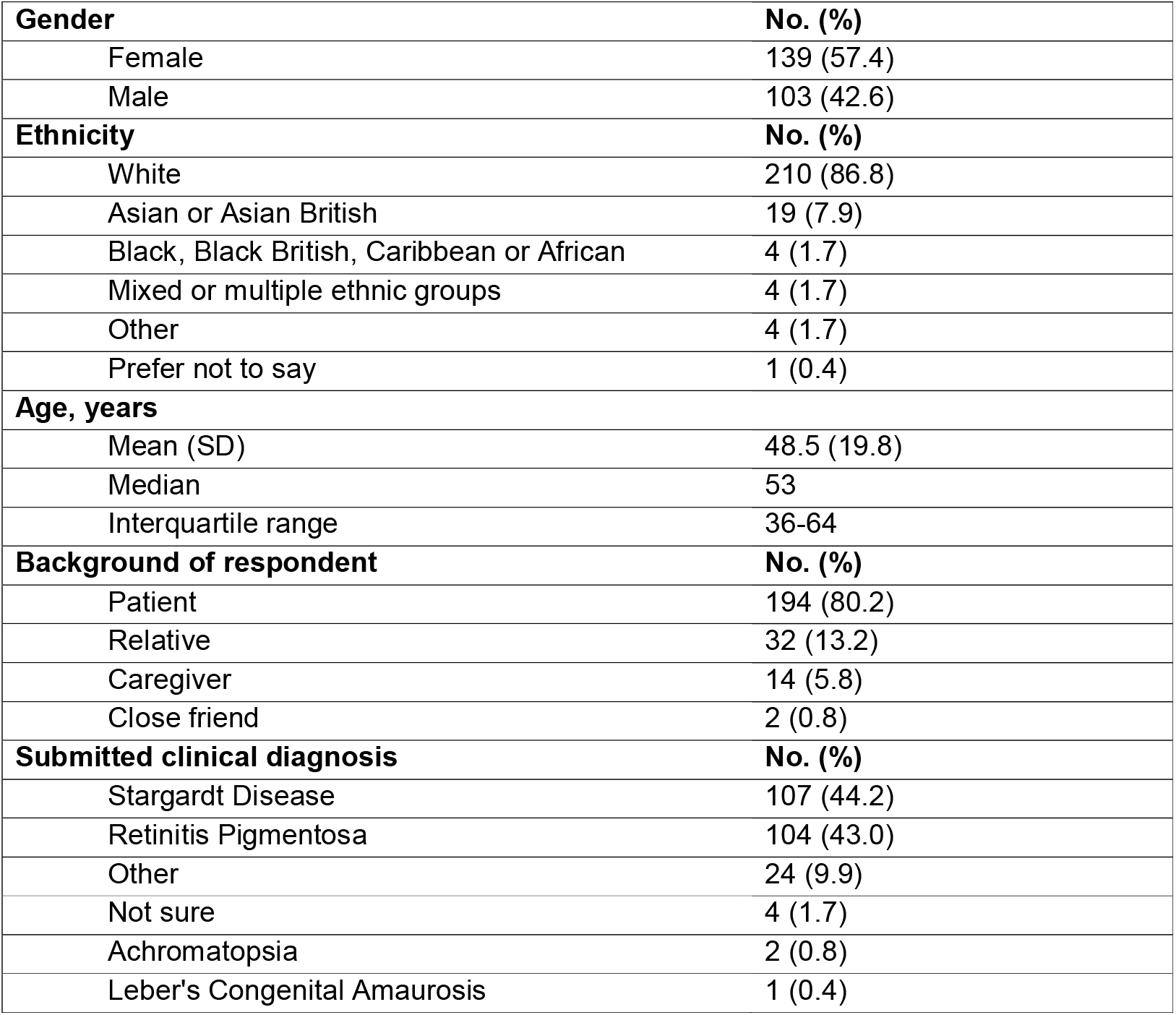
Demographics and characteristics of survey respondents.

| Demographics and Characteristics |
| --- |

|  |  |
| --- | --- |
| **Gender** | **No. (%)** |
| Female | 139 (57.4) |
| Male | 103 (42.6) |
| **Ethnicity** | **No. (%)** |
| White | 210 (86.8) |
| Asian or Asian British | 19 (7.9) |
| Black, Black British, Caribbean or African | 4 (1.7) |
| Mixed or multiple ethnic groups | 4 (1.7) |
| Other | 4 (1.7) |
| Prefer not to say | 1 (0.4) |
| **Age, years** |  |
| Mean (SD) | 48.5 (19.8) |
| Median | 53 |
| Interquartile range | 36-64 |
| **Background of respondent** | **No. (%)** |
| Patient | 194 (80.2) |
| Relative | 32 (13.2) |
| Caregiver | 14 (5.8) |
| Close friend | 2 (0.8) |
| **Submitted clinical diagnosis** | **No. (%)** |
| Stargardt Disease | 107 (44.2) |
| Retinitis Pigmentosa | 104 (43.0) |
| Other | 24 (9.9) |
| Not sure | 4 (1.7) |
| Achromatopsia | 2 (0.8) |
| Leber's Congenital Amaurosis | 1 (0.4) |

### Diagnostic journey

There was significant variability in the number of consultations required to reach a diagnosis, ranging from 1 to 50, with a median of 3 [interquartile range (IQR): 2-4]. It was observed that two respondents had indicated achromatopsia as the underlying IRD, and both reported that more than 10 consultations were required to reach a diagnosis. Similarly, the number of years to be seen by a specialist and the distance travelled to see a specialist varied widely, with a median of 1 year (IQR: 1-4) and 25 miles (IQR: 10 to 75), respectively (**Table 2**).

**Table 2.** Diagnostic experience.

| Diagnostic experience |  |
| --- | --- |
| No. of consultations for diagnosis |  |
| Mean (SD) | 3.7 (5.9) |
| Median | 3 |
| Interquartile range | 2-4 |
| No. of years to see a specialist |  |
| Mean (SD) | 5.2 (11.0) |
| Median | 1 |
| Interquartile range | 1 to 4 |
| Travelling distance to see specialist, miles |  |
| Mean (SD) | 73.9 (123.1) |
| Median | 25 |
| Interquartile range | 10 to 74 |
| Received a change in diagnosis |  |
| Yes | 88 (35.8%) |
| No | 158 (64.2%) |
| Offered a genetic test |  |
| Yes |  |
| Proceeded with test | 186 (75.6%) |
| Did not proceed with test | 5 (2.0%) |
| No | 55 (22.4%) |
| Genetic counselling before testing |  |
| Yes | 103 (41.9%) |
| No | 83 (33.7%) |
| Nil response | 60 (24.4%) |
| Year of genetic testing |  |
| 1980 to 1999 | 15 (6.1%) |
| 2000 to 2009 | 19 (7.7%) |
| 2010 to 2019 | 88 (35.8%) |
| 2020 to 2024 | 68 (27.6%) |
| Not applicable | 61 (24.8%) |
| Age at receiving results of genetic test |  |
| Mean (SD) | 47.5 (19.2) |
| Median | 50 |
| Interquartile range | 38 to 58 |

The diagnosis was revised in over a third of patients (35.8%). Not all patients were offered genetic testing (22.4%), but amongst those who were offered testing (77.6%), only a small percentage declined (2.6%, 5 out of 191). The majority who underwent genetic testing (72.4%) had the test performed before 2020. The median age was 42 years (IQR: 21 to 54) at the time they received the results of the genetic testing. The two most frequent diagnoses were Stargardt disease and RP for which the median age at diagnosis was 31 years (IQR: 15-48) and 50 years (IQR: 38-58), respectively.

### Communication of genetic testing and results

One-third of respondents did not undergo or recollected undergoing counselling prior to genetic testing (**Table 3**). Genetic test results were most frequently communicated verbally (in person, over the phone or via teleconference) in 39.9% of cases, followed by written correspondence (letter or electronic mail) in 27.3%. Where the results were communicated during an appointment, the median length of time was 20 minutes (IQR: 0 to 30 minutes).

**Table 3.** Patient communication and information needs.

| **Patient communication and information needs** |  |
| --- | --- |
| **Genetic counselling before testing** | **No. (%)** |
| Yes | 103 (56.0%) |
| No | 81 (44.0%) |
| **Communication of test results** | **No. (%)** |
| In-person | 70 (38.0%) |
| Letter | 62 (33.7%) |
| Over the phone | 18 (9.8%) |
| Via teleconference | 9 (4.9%) |
| Email | 8 (4.3%) |
| Results not received | 5 (3.3%) |
| Don't remember | 4 (2.2%) |
| No response | 8 (4.3%) |
| **Length of appointment for communication of results (minutes)** |  |
| Mean (SD) | 23.1 (24.4) |
| Median | 20 |
| Interquartile range | 0 to 30 |
| **Perception on understanding of results** | **No. (%)** |
| Definitely yes | 81 (33.5%) |
| Probably yes | 47 (19.4%) |
| Might or might not | 16 (6.6%) |
| Probably not | 23 (9.5%) |
| Definitely not | 17 (6.7%) |
| Not applicable | 58 |
| **Perception if questions were adequately answered** | **No. (%)** |
| Definitely yes | 56 (23.1%) |
| Probably yes | 52 (21.5%) |
| Might or might not | 39 (16.1%) |
| Probably not | 20 (8.3%) |
| Definitely not | 17 (7.0%) |
| Not applicable | 58 |
| **Preferences on being informed of progression quantitatively not qualitatively** |  |
| Prefer a great deal | 112 (46.3%) |
| Prefer a lot | 54 (22.3%) |
| Prefer a moderate amount | 42 (17.4%) |
| Prefer slightly | 22 (9.1%) |
| Do not prefer | 12 (5.0%) |

About half of the respondents indicated that they probably or definitely understood the results (52.8%), and a smaller proportion indicated that they definitely or probably felt their questions had been adequately answered (44.3%).

A thematic analysis of respondents’ replies regarding information they seek during follow-up visits revealed these to be the most frequently mentioned topics: treatment (35 responses); research (30 responses) or trials (20 responses); progression or deterioration (29 responses) or prognosis (16 responses) and family (11 responses).

### Attitude toward imaging-based and AI-augmented diagnosis

The vast majority of respondents indicated that they find it acceptable (80.6%) or somewhat acceptable (14.0%) to use ocular imaging to assess the likelihood of an IRD. Only a small percentage (1.6%) indicated they would find this unacceptable or somewhat unacceptable (**Table 4**).

**Table 4.** Patient perception on imaging and incorporation of AI.

| Patient perception |  |
| --- | --- |
| Preliminary assessment of status using an eye scan | No. (%) |
| Acceptable | 195 (80.6%) |
| Somewhat acceptable | 34 (14.0%) |
| Neither unacceptable nor acceptable | 9 (3.7%) |
| Somewhat unacceptable | 2 (0.8%) |
| Unacceptable | 2 (0.8%) |
| Perceived benefit of Eye2Gene AI for diagnosis | No. (%) |
| Strongly agree | 171 (70.7%) |
| Somewhat agree | 47 (19.4%) |
| Neither agree nor disagree | 19 (7.9%) |
| Somewhat disagree | 1 (0.4%) |
| Strongly disagree | 4 (1.7%) |
| Preference on access to undergoing retinal scans | No. (%) |
| Definitely yes | 158 (65.3%) |
| Probably yes | 57 (23.6%) |
| Might or might not | 18 (7.4%) |
| Probably not | 7 (2.9%) |
| Definitely not | 2 (0.8%) |
| Preference for local retinal scans to be sent to hospital | No. (%) |
| Definitely yes | 124 (51.2%) |
| Probably yes | 67 (27.7%) |
| Might or might not | 27 (11.2%) |
| Probably not | 15 (6.2%) |
| Definitely not | 9 (3.7%) |
| **Perceived acceptability of doctors using Eye2Gene** | **No. (%)** |
| Acceptable | 179 (74.0%) |
| Somewhat acceptable | 45 (18.6%) |
| Neither unacceptable nor acceptable | 11 (4.5%) |
| Unacceptable | 4 (1.7%) |
| Somewhat unacceptable | 3 (1.2%) |

A majority of respondents strongly agreed (70.7%) or somewhat agreed (19.4%) that incorporating AI would be beneficial if it can help clinicians reach the right diagnosis earlier; only a small proportion of respondents (2.1%) expressed disagreement.

With regards to access to undergoing imaging-based tests, most respondents expressed that they were in favour, with 65.4% indicating that they preferred access, an additional 23.6% expressed that they probably preferred access, and only a small proportion (3.7%) responded that they probably or definitely would not want access.

## Data Availability

All data produced in the present work are contained in the manuscript

## Discussion

The IRD pathway in the UK is highly variable, meaning that patients can experience significantly different diagnostic journeys depending on the health care provisions available locally to them. These differences include the waiting time to see a specialist (IQR 1 to 4 years), the commute involved to see a specialist (IQR 10 to 74 miles) and the number of visits needed to obtain a diagnosis (IQR 2 to 4). Furthermore, a substantial proportion of patients (35.8%) were initially given a diagnosis that was later changed. These findings are consistent with the previous literature investigating the diagnostic process of patients with IRDs [8,18]. Incorporating AI techniques, such as Eye2Gene (an AI tool currently under development for IRDs), that are capable of assessing retinal images for features of IRD can help streamline the diagnostic process, support clinical decision-making, and also improve patient access to expertise in IRD and thus standardise healthcare delivery [15].

Survey respondents demonstrated a positive attitude towards AI, with a clear majority (90.1%) receptive to its incorporation into the diagnostic testing of IRDs, especially if it assists the clinician in reaching the correct diagnosis more quickly (92.6%). These results are useful for several stakeholders, including healthcare providers, AI developers and regulatory bodies, as they inform the planning of implementation of AI in the care of patients with IRDs. Although the implementation of AI in healthcare remains in nascent stages, it is imperative that clinicians are trained about the principles and limitations of AI to ensure that AI is used responsibly.

This survey identified several topics frequently of interest to patients during consultations, offering invaluable insight for developing, implementing, evaluating, and improving AI tools intended for clinicians managing IRDs or patients affected by these conditions. Several topics are centred on the possibilities of treatment, including trials and research. The genetic heterogeneity of IRDs, coupled with rapid advances for both gene-specific and gene-agnostic approaches, generates a substantial body of knowledge that is challenging for clinicians to review efficiently and remain updated [19]. Supporting this point, the study revealed only 44.3% of patients were satisfied that their questions had been adequately addressed. Progress in AI and natural language processing may provide the means to achieving personalised, evidence-based medicine in IRD clinics by efficiently informing clinicians of a real-time summary of the literature relevant to the particular genetic diagnosis [20]. The accuracy and feasibility of using AI-based models to generate a summary of the scientific literature has been investigated, but not specifically for IRDs [20–22].

This survey additionally found that patients are not only keen to understand the disease trajectory but would also often prefer quantitative measures of progression instead of vague descriptions of deterioration. Objective markers of disease progression, such as expansion of areas of retinal pigment epithelial atrophy or increase in the extent of ellipsoid zone loss can be measured, but this manually laborious and time-consuming process is often limited to research studies with dedicated graders [22]. However, novel AI approaches may make it possible for clinicians to efficiently chart changes in these key disease features from clinical investigations, allowing for quantitative monitoring of disease progression [23]. Furthermore, the advent of wearable devices which collect detailed information on the visual behaviour of the patient, such as the distance at performing visual activities, vision duration and breaks, illumination level, and head movements, could greatly enrich and complement the structural insights provided by ocular imaging, offering a more comprehensive basis for disease prognostication [24].

To our knowledge, this survey is the first to evaluate patient perspectives and considerations regarding the application of AI in the diagnosis of IRDs. This survey includes a diverse range of patient demographics by ethnicity, age and genetic diagnosis, as well as representation from family members, caregivers and close friends.

There are several limitations to this study. First, the survey was only distributed electronically. This may introduce a sampling bias which favours individuals who are more inclined to adopt technological advances. Second, as this survey was distributed through UK-based charities, these findings may not be generalisable to other healthcare systems and regions outside the UK. Finally, recall bias may be present, as a substantial proportion of patients (72.4%) underwent genetic testing before 2020.

This study demonstrates substantial variations in the time it takes to obtain a diagnosis and the way that information is delivered and understood by patients evaluated in this study. AI algorithms such as Eye2Gene have the potential to address these unmet needs and potentially standardise the pathway to offer more patients equitable management and care. The majority of the patients, as well as their companions, have a positive attitude towards AI being incorporated to improve their clinical care in the UK. These findings, which reflect the perspectives of patients, are valuable for informing and shaping policy on the integration of AI into the IRD care pathway.

## Footnotes

**X:** @npontikos

WW and DS contributed equally. MM and NP contributed equally.

## Contributors

NP and MM conceived and designed the study. WW, DS and TM collected, analysed and interpreted data. WW, DS, MG, BT, CH, WAW, SA and LL co-designed the study. NP, MM, WW and DS wrote the manuscript. All authors contributed to critically reviewing the manuscript.

## Disclaimer

The funder had no role in study design, data collection, data analysis, data interpretation or writing of the report.

